# Oscillations of pause-burst neurons in the STN correlate with the severity of motor signs in Parkinson’s disease

**DOI:** 10.1101/2021.12.11.21267521

**Authors:** Elena M. Belova, Veronika I. Filyushkina, Indiko Dzhalagoniia, Anna A. Gamaleya, Alexey A. Tomskiy, Wolf-Julian Neumann, Alexey Sedov

## Abstract

**Background:** Oscillatory activity in the subthalamic nucleus (STN) in Parkinson’s disease (PD) is under extensive study. While rhythmic features of local field potentials are implicated in the manifestation of PD motor signs, less is known about single unit activity (SUA). SUA parameters inside the STN show significant heterogeneity, and various firing patterns may contribute unequally to PD pathophysiology.

**Objectives:** We searched for correlations between SUA parameters and PD motor signs, taking neuronal activity patterns into account.

**Methods:** 829 spike trains for STN SUA were recorded during 25 DBS surgeries. We have isolated three firing patterns (tonic, irregular burst and pause-burst) and, using mixed linear models, examined several ISI parameters and burst descriptors (for the last two patterns) for their correlation with the UPDRS 3 score and bradykinesia and rigidity scores on the contralateral body side.

**Results:** The predominance of pause-burst as opposed to tonic activity was associated with an increase in UPDRS 3 score. Oscillation scores in the alpha range correlated with bradykinesia and rigidity scores, and oscillation scores in the beta range correlated with bradykinesia score only for pause-burst neurons, while other patterns showed no correlation with PD motor signs. There was also significant negative correlation between bradykinesia score and theta oscillations for pause-burst pattern.

**Conclusions:** Pause-burst pattern and rhythmic neurons oscillating in the alpha range may affect motor processing in the basal ganglia more prominently than other activity patterns, probably reflecting progressive switching from tonic to burst to rhythmic activity in the parkinsonian state.

## 1. Introduction

Parkinson’s disease (PD) is a movement disorder, accompanied with dysfunction in the basal ganglia circuit communication, manifesting in pathological neuronal activity patterns. Levodopa administration is the most common therapy for PD, but in the case of emerging drug-induced motor complications, deep brain stimulation (DBS) is an effective treatment alternative. In this case, DBS electrodes are implanted in the basal ganglia nuclei, typically the subthalamic nucleus (STN). To validate the implant trajectory, intraoperative microelectrode recordings (MER) can support the definition of the exact borders of the target nuclei. This procedure provides unique access to neuronal activity in affected subcortical brain structures directly affected by PD pathophysiology. Therefore, deep brain recordings can facilitate the search for specific features associated with disease severity, or may reveal electrophysiological parameters predicting the optimal site for electrode implantation in order to support a good clinical outcome.

The most prominent motor signs of parkinsonism are tremor, bradykinesia and rigidity. Commonly, excessive neuronal oscillations in the basal ganglia network are implicated in the manifestation of clinical signs of parkinsonism. Tremor cells oscillating in the theta range are implicated in parkinsonian tremor, while bradykinesia and rigidity are attributed to the excessive oscillations in the basal ganglia network within the beta range (traditionally assigned as 13–30 Hz, but often varied in the exact thresholds) (1). Beta oscillations are thought to be the major manifestation of hypokinetic symptoms in the basal ganglia nuclei, as levodopa administration can significantly reduce both hypokinetic symptoms and oscillation power in the beta range within the STN (2).

Previously, we have shown heterogeneity of beta oscillations in local field potentials (LFPs) within the STN (1). Our findings suggest the existence of several sub-ranges of oscillations within 8–30 Hz borders, with alpha-beta oscillations associated with the bradykinesia scores and appearing to be partially insensitive to motor tests, whereas low and high beta oscillations were modulated in opposing ways during voluntary movements. This non-uniformity in beta oscillations may be also present in the STN single unit activity (SUA); however, this assumption has not been tested previously.

Observed SUA inside the STN displays obvious heterogeneity: spike trains with comparable discharge rate may be arranged evenly (tonic activity) or gathered into dense groups of spikes separated with sparse discharges or pauses (burst activity). The detection and isolation of distinct SUA patterns and the subsequent examination of their features has proved to be an effective approach to reveal particular parameters associated with the severity of movement disorders or particular clinical signs of the disease. Using this approach, several studies have reported that pauser cells activity parameters within the globus pallidus contribute to a particular manifestation of disease in dystonic patients (3–5). Previously, splitting STN neuronal activity into several patterns was used to examine the differences between parkinsonian and non-parkinsonian states (6), or to determine the spatial heterogeneity of SUA within the STN (7,8), probably reflecting STN functional topography and segregation between sensorimotor, limbic and associative regions (9). However, the association between clinical signs of PD and parameters of SUA for isolated neuronal patterns within the STN remains poorly understood.

In this study, we attempt to investigate features of STN SUA and their association with motor signs of PD using a large number of isolated single units and taking patterns of STN SUA into consideration. To isolate neuronal patterns, we have applied a previously developed approach utilizing unsupervised hierarchical clustering of spike trains based on spike density histograms (SDHs) (10). SDHs are particularly sensitive to pauses in spike trains, with pause number and duration affecting the ratio of the first two columns in SDHs, allowing better resolution for single unit bursts. This method deals with the issues related to poor specification of burst neurons and biased approaches to isolate firing patterns and allows the grouping of spike trains based on the inner structure and peculiarities of a given dataset.

## 2. Methods

### Patients

Twenty-five patients with mixed or akinetic-rigid subtypes of Parkinson’s disease participated in the study. All patients underwent DBS surgery to implant leads into the motor area of the subthalamic nucleus in 2016–2020 at the N.N. Burdenko Centre for Neurosurgery. One day before the surgery, patients underwent a neurological examination using Unified Parkinson’s Disease Rating Scale (UPDRS) to assess motor symptoms of PD on both sides of the body after 12 hours without medications (off-state). In addition to a total score for motor symptoms (UPDRS III), severity of rigidity (item 22) and bradykinesia (items 23-26) on the contralateral side of the body were assessed and accounted separately.

The details for all procedures were carefully explained to the participants before the surgery. All participants gave written informed consent prior to inclusion in the study. The study protocol was approved by the local Ethics Committee, in agreement with the Declaration of Helsinki. Clinical data for the patients are shown in Table 1.

**Table 1.** Summary of clinical data for PD patients included in the study

| patient | DBS<br>sides | start age,<br>yrs | age, yrs | PD<br>duration,<br>yrs | UPDRS3<br>off score | bradykinesia<br>off score (L/R) | rigidity<br>off score<br>(L/R) | tremor<br>off score<br>(L/R) |
| --- | --- | --- | --- | --- | --- | --- | --- | --- |
| 1 | L+R | 40-44 | 50-54 | 11 | 62 | 10/13 | 6/4 | 5/3 |
| 2 | L+R | 30-34 | 50-54 | 20 | 56 | 12/14 | 6/4 | 0/0 |
| 3 | L+R | 50-54 | 60-64 | 9 | 28 | 4/7 | 1/2 | 0/2 |
| 4 | L+R | 35-39 | 45-49 | 9 | 51 | 9/8 | 6/6 | 5/2 |
| 5 | L+R | 35-39 | 45-49 | 12 | 57 | 12/10 | 6/4 | 5/5 |
| 6 | L | 30-34 | 35-39 | 5 | 37 | 12 | 6 | 7 |
| 7 | L+R | 35-39 | 45-49 | 14 | 41 | 7/10 | 4/4 | 7/2 |
| 8 | L+R | 55-59 | 60-64 | 8 | 52 | 11/10 | 3/4 | 0/2 |
| 9 | L+R | 45-49 | 60-64 | 13 | 41 | 6/6 | 3/3 | 2/3 |
| 10 | L+R | 35-39 | 50-54 | 12 | 78 | 14/11 | 8/6 | 7/2 |
| 11 | L+R | 30-34 | 40-44 | 9 | 37 | 4/3 | 3/2 | 7/6 |
| 12 | L+R | 25-29 | 35-39 | 10 | 36 | 7/6 | 5/4 | 6/0 |
| 13 | L+R | 35-39 | 60-64 | 22 | 54 | 12/8 | 6/4 | 2/1 |
| 14 | R | 50-54 | 60-64 | 11 | 44 | 5 | 5 | 0 |
| 15 | L+R | 40-44 | 50-54 | 13 | 71 | 16/16 | 4/6 | 4/4 |
| 16 | L+R | 45-49 | 55-59 | 11 | 43 | 14/6 | 5/2 | 0/0 |
| 17 | L+R | 35-39 | 45-49 | 8 | 32 | 8/6 | 4/3 | 0/0 |
| 18 | L+R | 40-44 | 44-49 | 9 | 39 | 8/4 | 6/5 | 1/0 |
| 19 | L+R | 35-39 | 50-54 | 12 | 46 | 11/9 | 2/3 | 0/0 |
| 20 | L+R | 50-54 | 65-69 | 13 | 18 | 6/1 | 4/0 | 0/0 |
| 21 | L+R | 35-39 | 40-44 | 7 | 78 | 13/16 | 6/7 | 8/4 |
| 22 | L+R | 45-49 | 50-54 | 8 | 59 | 9/10 | 4/2 | 5/5 |
| 23 | L+R | 35-39 | 50-54 | 14 | 26 | 3/0 | 3/3 | 6/6 |
| 24 | L+R | 35-39 | 45-49 | 8 | 24 | 3/7 | 2/4 | 0/2 |
| 25 | L+R | 40-44 | 55-59 | 12 | 50 | 6/11 | 4/6 | 0/5 |

### Procedure and data collection

Data for spontaneous SUA were collected intraoperatively. Before the surgery, patients were instructed to withdraw all anti-parkinsonian medications overnight. MER were performed in the awake state under local anesthesia. Magnetic resonance imaging (MRI) was performed preoperatively, with a fixed stereotactic frame using T1 and T2 weighted sequences on a Signa Horizon Speed 1.5T scanner (General Electric); MRI scans and Leksell G Surgiplan Software (Elekta, Sweden) were used to calculate target coordinates, microelectrode recordings were performed to localize the borders of the STN and provide information for proper implantation for stimulating electrodes. The electrode was placed 10 mm above the target and advanced in 0.1–0.3 mm steps along the trajectory, using the NeuroNav system (AlphaOmega, Israel). Extracellular multiunit activity (up to 3–4 single units) was recorded by high resistance (0.5–1 MegaOhm) tungsten microelectrodes (NeuroProbe, AlphaOmega, Israel).

### Data processing

MER signals were pre-processed, including bandpass filtering (300–5000 Hz), alignment and spike sorting, with Spike2 software (CED, Cambridge, UK). Spikes were detected using an amplitude threshold and then sorted using principal component analysis (PCA). Records with 1–3 clearly identifiable single units (i.e. steady forms of spikes and amplitude differences more than 20% along the record, which were clearly recognized on the PCA plot) were selected for further analysis. We used the data for isolated single units with a good signal-to-noise ratio (more than 3 standard deviations from background noise range), containing more than 200 spikes and longer than 10 seconds. Recordings with an unclear number of single units were discarded. For each spike train, we calculated several parameters based on interspike intervals (ISIs), including regular ISI descriptors (firing rate, median, coefficient of variation, standard deviation of ISI) (11) and strength of neuronal oscillations using oscillation scores in several frequency bands (3–8, 8–12, 12–20, 20–30, 30–60 and 60–90 Hz), as described in (12). In brief, autocorrelation histogram (ACH) for a given spike train is computed, smoothed, and then the central peak is removed; power spectra of peakless ACH is calculated using fast Fourier transform. Oscillation scores for a given frequency band is calculated as a ratio of peak amplitude to mean spectral amplitude in the band (12).

In addition, spike train data were divided into three patterns based on similarities of the spike density histograms using a hierarchical clustering approach. Namely, the tonic pattern represents regular activity with similar intervals between the spikes close to the mean ISI; the irregular-burst pattern has varying ISIs without obvious prolonged pauses in spike trains; and the pause-burst pattern contains bursts of spikes separated by pauses in activity. Parameters of SUA for the three isolated patterns were analyzed separately.

For irregular-burst and pause-burst patterns of activity, we isolated bursts in spike trains and examined the features reflecting burst frequency and duration, and the density of spikes inside the bursts. Briefly, bursts in spike trains were detected using the Poisson surprise approach using S = 3, which compares the spike train and a random sequence of spikes following the Poisson distribution (13). Bursts are determined as sequences of spikes, where the number of spikes in a given interval significantly (i.e. more than 3 negative logarithms of Poisson probability) exceeds the occurrence rate of spikes for the Poisson distribution with the same firing rate. After burst detection, we calculated several parameters for isolated bursts, including intraburst and interburst intervals, mean and median ISI in the bursts, mean number of spikes in the bursts, mean duration of the bursts, and percentage of spikes in the bursts.

To examine properties of pause-burst single units with high rhythmic activity, we separated pause-burst single units with oscillation scores exceeding the 0.75 quantile in the 8–12, 12–20 and 20-30 Hz ranges in order to compare their parameters with the rest of the pause-burst single units.

### Statistical analysis

Statistical analysis of the data was performed using R (https://cran.r-project.org). Differences between the patterns were assessed using linear models for activity patterns as an ordered factor variable (tonic > irregular-burst > pause-burst). Prior to the model fitting, dependent variables were normalized and checked for normal distribution using the Lilliefors test for the composite hypothesis of normality (mean and variance unknown) (14). For non-normally distributed parameters, the Kruskal-Wallis test for independent samples was used (15).

The associations between clinical symptoms and parameters of SUA were analysed using mixed effect linear models, allowing us to account for non-independence of the data within each subject (16). We recorded multiple isolated spike trains for each hemisphere, representing a set of dependent data characterizing STN neuronal activity in a given hemisphere, whereas mixed effect models deal with pseudoreplication issues and allow processing of large samples with non-independent data. The R package “nlme” was used for calculations.

Prior to model fitting, dependent continuous variables were z-normalized and checked for normal distribution, and independent discrete variables representing the severity of parkinsonian symptoms were scaled to the [-2, 2] range, so the variance of variables were scaled to 1. Associations between rhythmic parameters of neuronal activity (oscillation scores in theta, alpha, low beta, high beta or gamma range were used as a dependent target variable) and motor signs of PD (UPDRS 3 off-score, rigidity or bradykinesia score were used as an independent variable) were assessed for all three patterns together and then for each pattern separately. In all cases, patient identifiers were used as a random factor to nest all spike trains recorded in this patient. Models for UPDRS III score, bradykinesia and rigidity scores were fitted separately. Association between oscillation scores and clinical signs were estimated using a slope coefficient for fixed effects part of the model output.

Differences between the features of non-oscillating units and units oscillating in the 8–30 Hz range were estimated using Student’s t-test. The association between clinical signs and the predominance of oscillating and non-oscillating single units was assessed using the Kruskal-Wallis rank sum test.

P-values obtained during statistical analysis were adjusted for multiple comparisons using the false discovery rate (FDR) approach. Adjusted p values <0.05 were accepted as statistically significant. Values are shown as the median with interquartile range (IQR) in parentheses, unless otherwise specified.

## 3. Results

### Patterns of activity inside the STN

Overall, 829 single units from 48 hemispheres were assigned to the dataset. For each patient, 15–89 single units were isolated (median number 29), median duration of spike trains was 16.9 s (13.6–22.9 s) containing 609 spikes on average (400–938). Spike trains from the whole dataset were grouped into three patterns—tonic, irregular-burst and pause-burst—based on normalized spike density histograms and hierarchical clustering (Figure 1a). Tonic neurons displayed regular activity with comparable intervals between the spikes (Figure 1b), irregular-burst neurons were characterized by irregular intervals between the spikes and emerging spike bursts without prominent silence periods in spike trains (Figure 1c), whereas pause-burst neurons appeared to have obvious silent intervals separating periods of high frequency discharges (Figures 1d, e).

**Figure 1.**
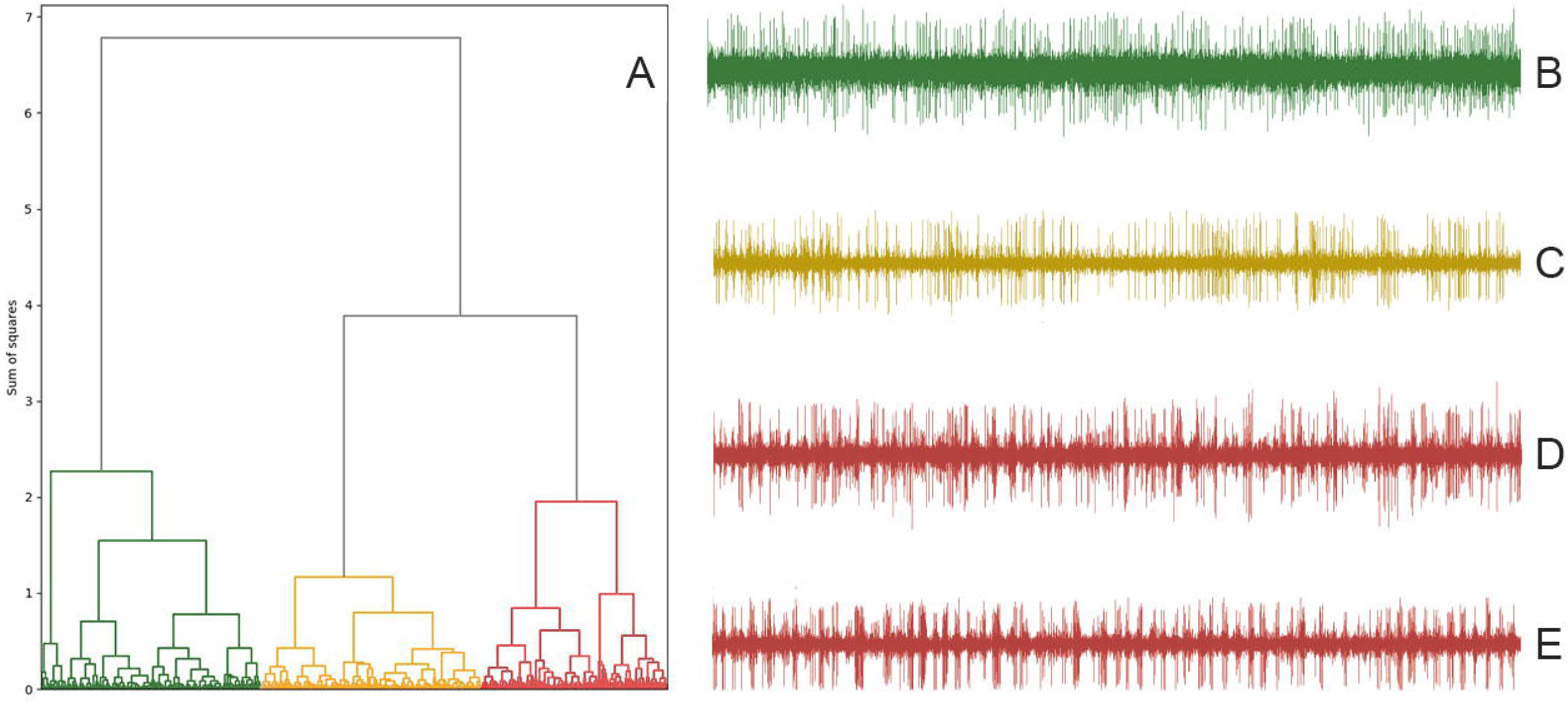
Clustering dendrogram based on spike density histograms (A) and example raw recordings for spike trains with tonic (B), irregular-burst (C) and non-rhythmic (D) and rhythmic (E) pause-burst patterns of single unit activity inside the STN.

Pattern prevalence inside the nucleus varied from 4.2 to 69.7% for the tonic pattern (total count of cells recorded was 283), from 17.9 to 57.1% for the irregular-burst pattern (total count = 293), and from 3.0 to 52.6% for the pause-burst pattern (total count = 253). Spike trains within the patterns differed significantly in terms of ISI descriptors affecting spike density histograms and the corresponding position inside the clustering dendrogram, i.e. CV ISI (p-value = 2.2 × 10^−16^) and asymmetry index (p-value = 2.2 × 10^−16^).

Additionally, we found significant differences in firing rates (median values for tonic, irregular-burst and pause-burst single units were 41.9, 35.8 and 28.3 spikes/s, respectively, p-value =1.98 × 10^−13^). There was also significant heterogeneity in pattern prevalence along the electrode trajectory inside the STN: pause-burst single units were shifted towards the dorsal half of the STN, whereas tonic units were prevalent in the ventral part of the STN (p-value = 2 × 10^−16^). A summary of the parameters of neuronal activity for the three isolated patterns is shown in Table 2.

**Table 2.**
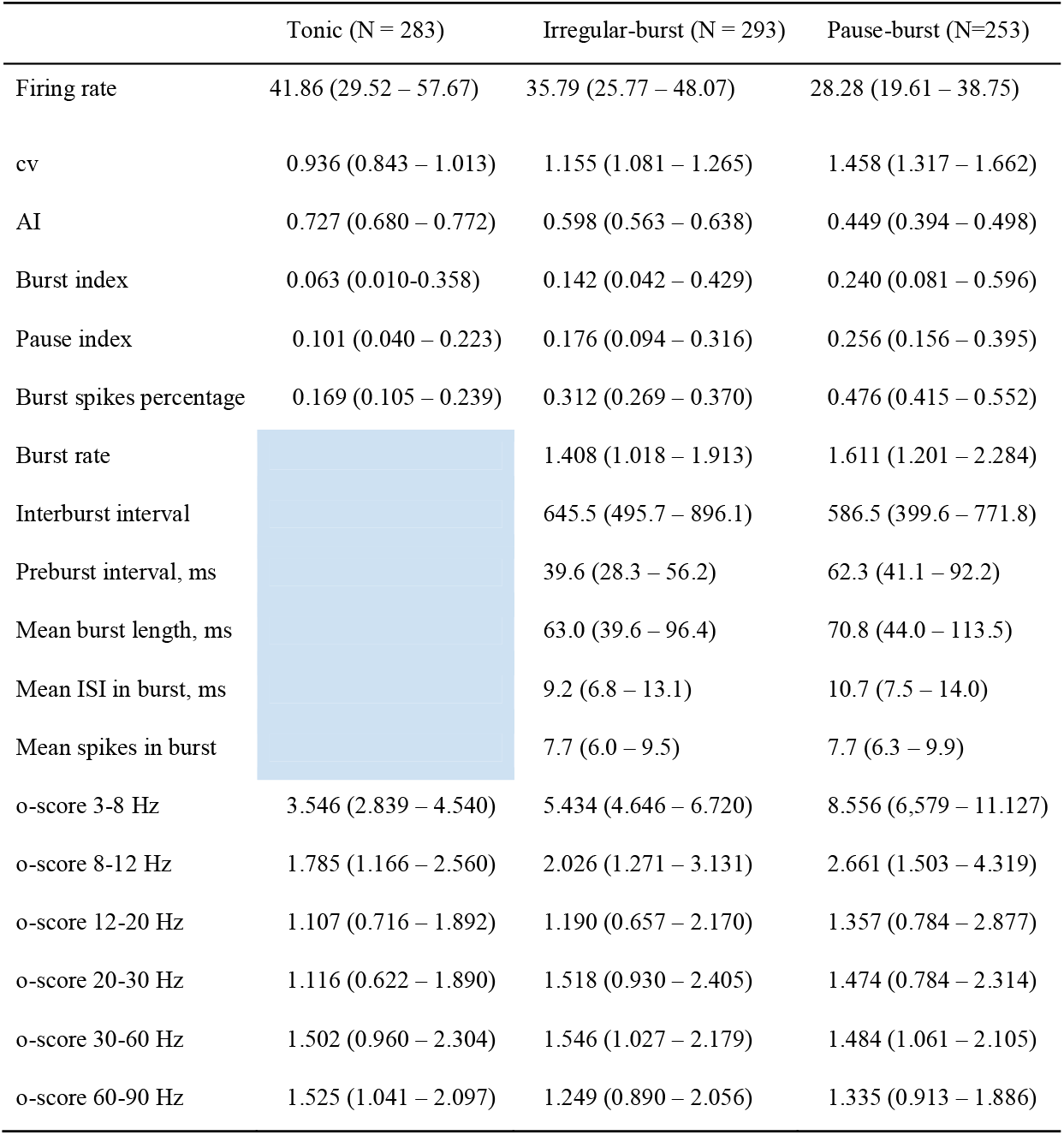
Median parameter values with IQR (in parentheses) for three isolated patterns of single unit activity in the STN

### Correlation between SUA patterns, rhythmic parameters and clinical signs of PD

We examined the association between parkinsonian motor symptoms (UPDRS 3 score in the off-state, and bradykinesia and rigidity scores on the contralateral side of the body) and neuronal activity parameters inside the STN and the predominance of isolated patterns. First of all, we found a significant correlation between pattern prevalence and UPDRS 3 score in the off-state, with a higher percentage of pause-burst neurons associated with a higher UPDRS 3 score (p-value = 0.015) (Figure 2).

**Figure 2.**
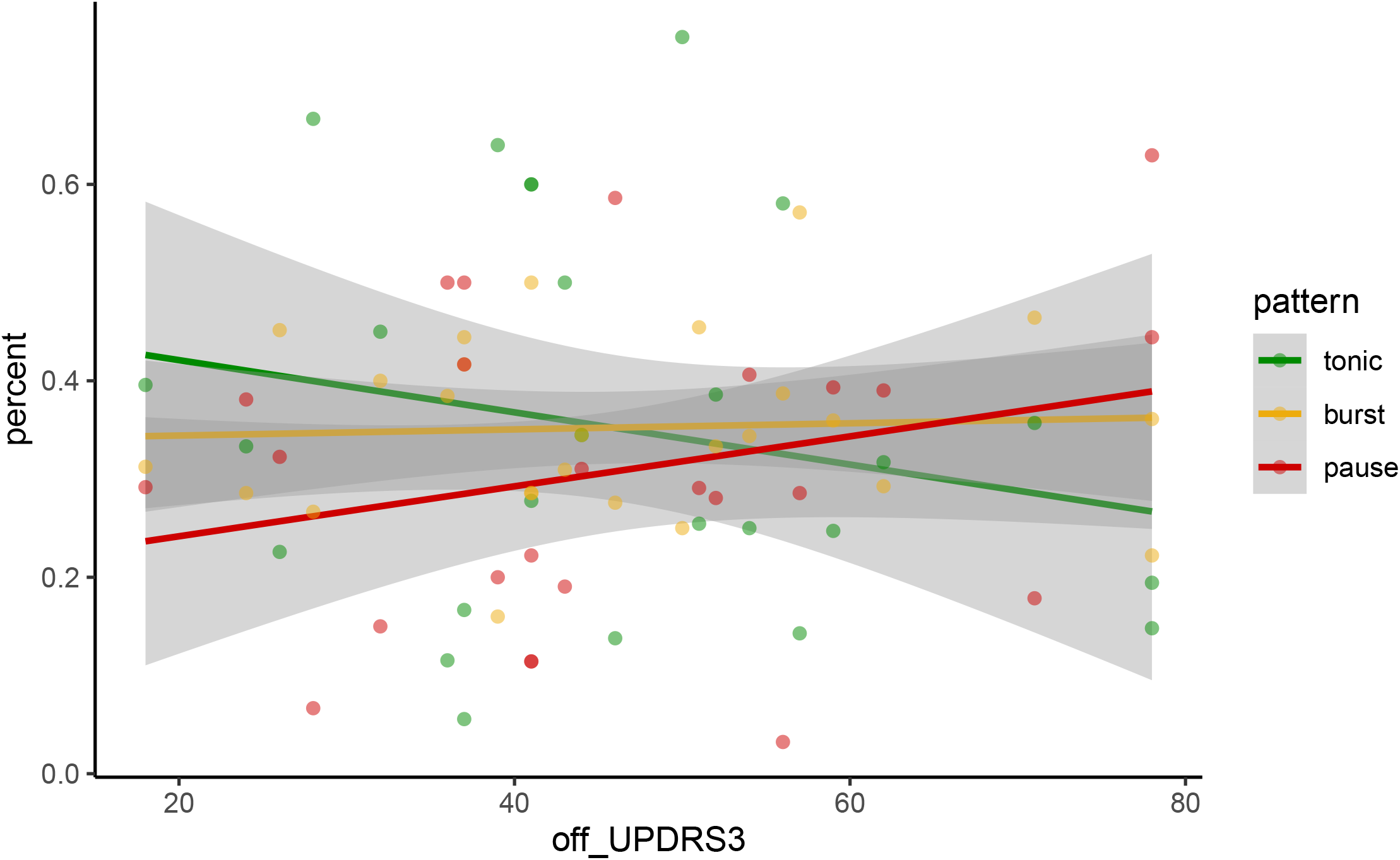
Association between pattern prevalence inside the STN and UPDRS 3 off score

For the whole spike trains dataset, we found significant correlation between UPDRS 3 scores in the off-state and oscillation scores in alpha range (8-12 Hz, adjusted p-value = 0.0454). When searching for associations between specific motor signs of PD and oscillation scores, alpha oscillations again was the only range that correlated significantly with bradykinesia and rigidity on the contralateral side of the body (FDR adjusted p-values for mixed linear models were 0.009 and 0.0007, respectively) Regression slopes and adjusted p-values for linear mixed effect model fitting are shown in Table 3.

**Table 3.** Slopes for association between oscillation scores and parkinsonian motor signs (UPDRS 3 score, and bradykinesia and rigidity scores on contralateral side of the body) estimated by mixed effect linear models

|  | **All_patterns** | **Tonic** | **Irregular burst** | **Pause burst** |
| --- | --- | --- | --- | --- |
| UPDRS3 off-score | Th -0.0104<br>p-value =0.88<br>**AI 0.2209\***<br>**p-value =0.045**<br>LB 0.1627<br>p-value =0.098<br>HB 0.089<br>p-value =0.098<br>Gm -0.0207<br>p-value =0.88 | Th 0.0203<br>p-value =0.8189<br>AI 0.0383<br>p-value =0.7089<br>LB -0.0566<br>p-value =0.7089<br>HB 0.0041<br>p-value =0.9489<br>Gm -0.127<br>p-value =0.4657 | Th -0.0443<br>p-value =0.45<br>AI 0.1238<br>p-value =0.45<br>LB 0.0742<br>p-value =0.45<br>HB 0.05<br>p-value =0.45<br>Gm 0.0572<br>p-value =0.45 | Th -0.1369<br>p-value =0.08<br>**AI 0.3531 \***<br>**p-value =0.015**<br>**LB 0.3272 \***<br>**p-value =0.015**<br>**HB 0.161 \***<br>**p-value =0.015**<br>Gm 0.0042<br>p-value =0.95 |
| Bradykinesia score (both sides) | Th -0.0192<br>p-value =0.89<br>**AI 0.1794\*\***<br>**p-value =0.009**<br>LB 0.081<br>p-value =0.22<br>HB 0.085<br>p-value =0.08<br>Gm -6e-04<br>p-value =0.99 | Th -0.0303<br>p-value =0.77<br>AI -0.0228<br>p-value =0.77<br>LB -0.0498<br>p-value =0.77<br>HB 0.0163<br>p-value =0.78<br>Gm -0.0832<br>p-value =0.77 | Th -0.0045<br>p-value =0.93<br>AI 0.1507<br>p-value =0.16<br>LB -0.0098<br>p-value =0.93<br>HB 0.0104<br>p-value =0.93<br>Gm 0.0058<br>p-value =0.93 | **Th -0.1946 \***<br>**p-value =0.01**<br>**AI 0.2895 \*\***<br>**p-value =0.006**<br>**LB 0.2531 \***<br>**p-value =0.01**<br>**HB 0.1914 \*\***<br>**p-value =0.0006**<br>Gm 0.0311<br>p-value =0.64 |
| Rigidity score (both sides) | Th 0.0792<br>p-value =0.11<br>**AI 0.1866**<br>**p-value =0.0007**<br>LB 0.1318<br>p-value =0.009<br>HB 0.0747<br>p-value =0.08<br>Gm 0.022<br>p-value =0.60 | Th 0.0691<br>p-value =0.54<br>AI 0.0348<br>p-value =0.99<br>LB 5e-04<br>p-value =0.99<br>HB 0.0251<br>p-value =0.99<br>Gm 0.0161<br>p-value =0.99 | Th 0.084<br>p-value =0.13<br>AI 0.1356<br>p-value =0.13<br>LB 0.0061<br>p-value =0.93<br>HB 0.0153<br>p-value =0.93<br>Gm 0.0176<br>p-value =0.93 | Th 0.0914<br>p-value =0.18<br>**AI 0.2555**<br>**p-value =0.01**<br>LB 0.1746<br>p-value =0.06<br>**HB 0.1401**<br>**p-value =0.01**<br>Gm 0.0219<br>p-value =0.72 |

When we examined the significance of association between UPDRS 3 score and rhythmic parameters within each pattern, the pause-burst pattern was the only SUA pattern possessing significant correlations with oscillation scores representing rhythmic features of activity. Oscillation scores in the alpha, low beta and high beta bands in pause-burst neurons correlated significantly with UPDRS 3 score (adjusted p-values were 0.015 for all three bands; see Table 3).

Then, we searched for associations between specific motor signs of PD and oscillation scores for each pattern of activity. We have found that only pause-burst single units revealed significant correlations with parkinsonian motor symptoms (Figure 3). We have found a significant positive association between oscillation scores in the 8–30 Hz range (comprising alpha, low beta and high beta ranges) for pause-burst neurons, and bradykinesia scores (adjusted p-values were 0.006, 0.01 and 0.0006, respectively) (Figure 3) and significant negative association with oscillations scores in the 3–8 Hz range (theta range) for pause-burst neurons and bradykinesia scores (adjusted p-value = 0.001). No significant association was found for either tonic or pause-burst patterns of activity (Figure 3). There were also positive correlation between rigidity score and alpha and high beta bands (adjusted p-values were 0.01). No significant correlation was found between motor signs on the contralateral side of the body and tonic and irregular burst patterns of activity.

**Figure 3.**
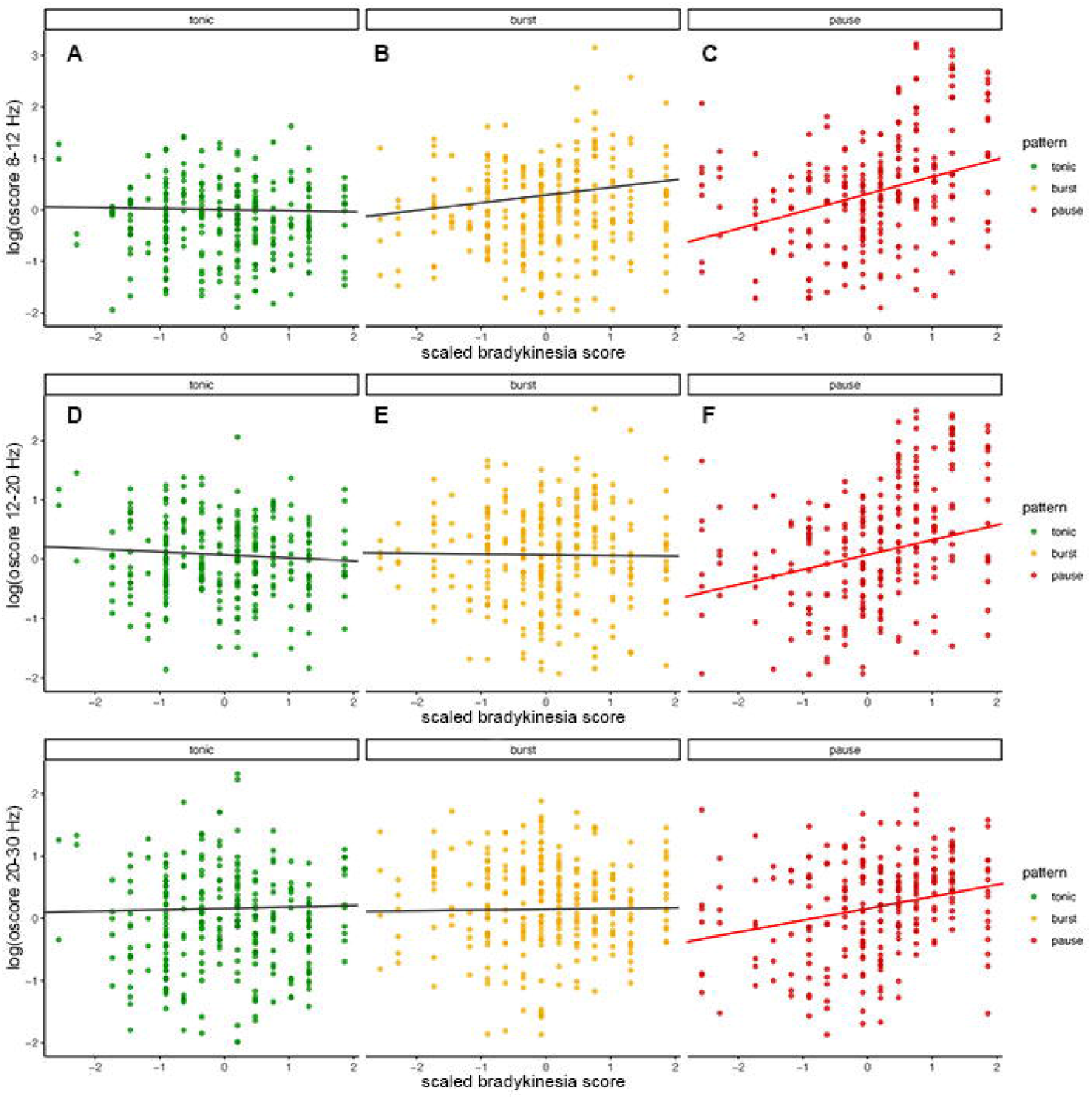
Association between oscillation scores in 8-12 (A,B,C), 12-20 (D,E,F) and 20-30 (G,H,I)Hz ranges and bradykinesia scores on the contralateral body side for tonic (A,D,G), irregular-burst (B,E,H,) and pause-burst (C,F,I) patterns of single unit activity inside the STN. Regression slopes with p-values less than 0.05 are shown in red

### The examination of pause-burst pattern of activity with high alpha-beta oscillation scores

For our data, oscillation scores for STN single units in the 8–12 and 12–20 Hz ranges correlate strongly (r = 0.889, p-value < 0.001), while there was only modest correlation between high beta and alpha or low beta range (r = 0.24 and 0.29, respectively, p-value < 0.001). Examination of several ISI parameters (firing rate, coefficient of ISI variation and asymmetry index) and burst descriptors corresponds well with this notion. Single units oscillating in either alpha, low or high beta ranges differ significantly from pause-burst units with weak oscillation scores in these ranges (designated further as non-oscillating units) (Table 4). However, coefficient of ISI variation was the single main parameter discriminating all alpha-beta oscillating units from non-oscillating units (p-value <0.01). Low beta oscillating units was the only group differed significantly from non-oscillating units in firing rate, while only alpha units differed significantly by asymmetry index (Table 4). However, when considering parameters of burst activity, alpha and low beta oscillating units both differed significantly from non-oscillating units by burst index, burst rate, interburst interval, mean burst length and median ISI in burst, while high beta oscillating units displayed significant difference from non-oscillating units in values of pause index and preburst interval (Table 4). All considered groups have comparable percent of spikes in bursts.

**Table 4.**
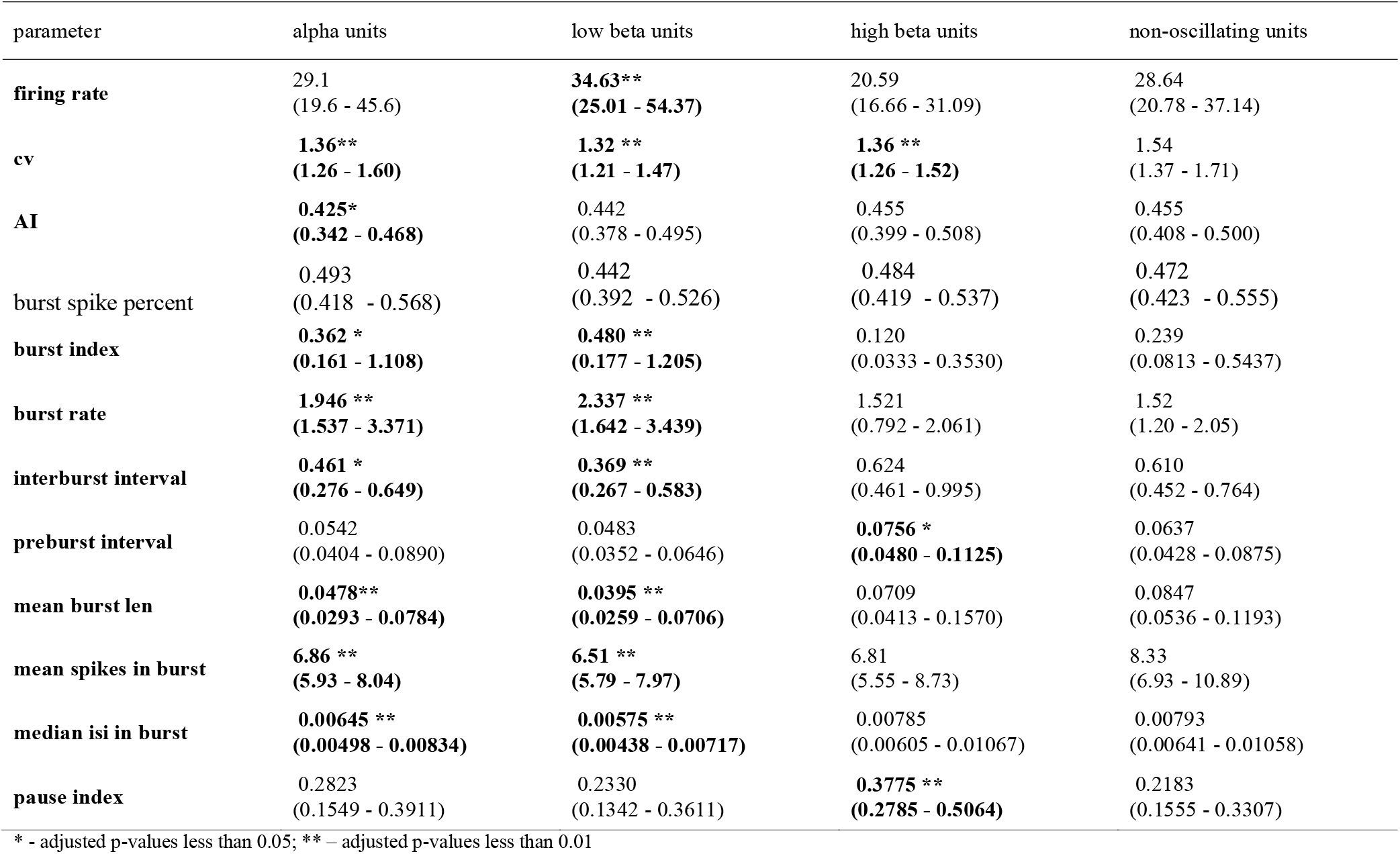
Median parameter values with IQR (in parentheses) for pause-burst units with high oscillation scores in 8-30 Hz range (alpha, low beta and high beta units) and low oscillation scores in 8-30 Hz (non-oscillating units).

| parameter | alpha units | low beta units | high beta units | non-oscillating units |
| --- | --- | --- | --- | --- |
| **firing rate** | 29.1<br>(19.6 - 45.6) | **34.63\*\***<br>**(25.01 - 54.37)** | 20.59<br>(16.66 - 31.09) | 28.64<br>(20.78 - 37.14) |
| **cv** | **1.36\*\***<br>**(1.26 - 1.60)** | **1.32 \*\***<br>**(1.21 - 1.47)** | **1.36 \*\***<br>**(1.26 - 1.52)** | 1.54<br>(1.37 - 1.71) |
| **AI** | **0.425\***<br>**(0.342 - 0.468)** | 0.442<br>(0.378 - 0.495) | 0.455<br>(0.399 - 0.508) | 0.455<br>(0.408 - 0.500) |
| burst spike percent | 0.493<br>(0.418 - 0.568) | 0.442<br>(0.392 - 0.526) | 0.484<br>(0.419 - 0.537) | 0.472<br>(0.423 - 0.555) |
| **burst index** | **0.362 \***<br>**(0.161 - 1.108)** | **0.480 \*\***<br>**(0.177 - 1.205)** | 0.120<br>(0.0333 - 0.3530) | 0.239<br>(0.0813 - 0.5437) |
| **burst rate** | **1.946 \*\***<br>**(1.537 - 3.371)** | **2.337 \*\***<br>**(1.642 - 3.439)** | 1.521<br>(0.792 - 2.061) | 1.52<br>(1.20 - 2.05) |
| **interburst interval** | **0.461 \***<br>**(0.276 - 0.649)** | **0.369 \*\***<br>**(0.267 - 0.583)** | 0.624<br>(0.461 - 0.995) | 0.610<br>(0.452 - 0.764) |
| **preburst interval** | 0.0542<br>(0.0404 - 0.0890) | 0.0483<br>(0.0352 - 0.0646) | **0.0756 \***<br>**(0.0480 - 0.1125)** | 0.0637<br>(0.0428 - 0.0875) |
| **mean burst len** | **0.0478\*\***<br>**(0.0293 - 0.0784)** | **0.0395 \*\***<br>**(0.0259 - 0.0706)** | 0.0709<br>(0.0413 - 0.1570) | 0.0847<br>(0.0536 - 0.1193) |
| **mean spikes in burst** | **6.86 \*\***<br>**(5.93 - 8.04)** | **6.51 \*\***<br>**(5.79 - 7.97)** | 6.81<br>(5.55 - 8.73) | 8.33<br>(6.93 - 10.89) |
| **median isi in burst** | **0.00645 \*\***<br>**(0.00498 - 0.00834)** | **0.00575 \*\***<br>**(0.00438 - 0.00717)** | 0.00785<br>(0.00605 - 0.01067) | 0.00793<br>(0.00641 - 0.01058) |
| **pause index** | 0.2823<br>(0.1549 - 0.3911) | 0.2330<br>(0.1342 - 0.3611) | **0.3775 \*\***<br>**(0.2785 - 0.5064)** | 0.2183<br>(0.1555 - 0.3307) |

\* - adjusted p-values less than 0.05; \*\* – adjusted p-values less than 0.01

## 4. Discussion

The present study demonstrates the association of parkinsonian motor signs with features of SUA patterns that can be isolated within the STN along the microelectrode trajectory. Using spike density histograms and a hierarchical clustering approach (10), we have performed unsupervised isolation of three patterns designated here as tonic, irregular-burst and pause-burst activity. We then searched for particular rhythmic features within each pattern, assuming that different patterns and oscillation bands may contribute unequally to motor control impairment within the basal ganglia network.

We have shown that the prevalence of tonic activity was associated with a lower UPDRS 3 score in the off-state compared to patients with predominant pause-burst SUA in the STN. In all recordings, tonic single units tend to occupy ventral areas of the STN, while pause-burst units are usually seen more dorsally. This corresponds well to the reports in the literature (7–9), with some authors suggesting that tonic activity represents associative or limbic areas that lie in the ventral regions of the STN (9). However, tonic activity may be also seen in the most dorsal parts of the STN, though this pattern is less frequent in the upper half of the microelectrode trajectory, which targets the caudal parts of the nucleus corresponding to the STN motor area.

Alpha oscillation score appeared to correlate with the severity of PD motor symptoms for the whole set of spike trains, independently of a particular activity pattern. However, only the pause burst pattern of activity revealed associations of several rhythmic features of spike trains with parkinsonian motor signs. Oscillations at alpha, low beta and high beta frequencies (8-12, 12–20 and 20-30 Hz) correlated with off-state UPDRS 3 score and specifically with the severity of bradykinesia on the contralateral body side, while oscillations in the alpha and high beta range (8–12 and 203-0 Hz) revealed an additional association with rigidity score contralaterally. These findings indicate that oscillations at various frequencies have unequal impact on the STN units when considering PD motor impairment. Presumably, excessive oscillations at lower frequencies inside broad alpha-beta band display higher impact on the neuronal activity in the STN, while oscillations at higher frequencies are less prominent and may be “masked” by the units exhibiting other discharge patterns. Also, we have found significant negative correlation between theta oscillation score and bradykinesia score for pause-burst pattern.

Previously, the most comprehensive study of SUA features in the STN and their association with motor signs of parkinsonism has been performed by Sharrott et al. (2014), who examined the correlation between the proportion of STN single units exhibiting certain activity features and the severity of parkinsonian motor symptoms (17). Although they revealed a correlation between rhythmic parameters of the STN single units and motor signs of PD (in particular, an association between percent of beta (14-31 Hz) oscillating neurons and limb rigidity score), they did not inspect patterns of SUA within the STN, despite the fact that their alterations may contribute significantly to PD pathophysiology.

Several inconsistencies of our data with the results of previous study of Sharrott et al. (2014) may be due to differences in details of the data analysis. Briefly, Sharrott et al. identified oscillating units using a spectral approach: they searched for spectra where the oscillating power of three contiguous bins in the range of interest exceeded the 95% confidence limit within the frequency range (based on 1000 times ISI shuffling). We have estimated rhythmic features of STN neurons using oscillation scores that allow quantitative evaluation of oscillations for the whole set of STN neurons. Another difference related to the oscillation ranges that were applied to search for oscillations: here we used a more comprehensive set of oscillations consisting of theta (3–8 Hz), alpha (8–12 Hz), and low (12–20 Hz) and high (20–30 Hz) beta and gamma (30–60 and 60–90 Hz) ranges, while the previous study dealt with broad sub-beta (3–13 Hz), beta (14–31 Hz) and gamma (40–90 Hz) ranges. This inconsistency in splitting oscillations into particular frequency ranges may also affect the findings. We also used linear mixed effect models, allowing us to nest all of the data recorded inside one subthalamic nucleus when searching for correlation with clinical symptoms instead of averaging the power or calculating the maximal power within each range and other computational procedures applied by Sharrott et al. (2014).

Oscillations in the alpha band appeared to be the only ones to correlate with PD motor signs when examining all SUA patterns. This may be due to higher stability of oscillations at lower frequencies, that may impair switching to another state required to normal signal processing through the basal ganglia. These findings agree with our previous research, showing that alpha-low beta oscillation power in STN LFP recordings increased with the bradykinesia score with the most intense association between the features found in the 8–12 Hz range, corresponding to alpha oscillations (1).

Among pause-burst cells, neurons oscillating in the broad alpha-beta range appeared to be tightly linked with parkinsonian symptoms. However, detailed analysis of activity features implied the existence of at least two distinct types of rhythmic activity related to either alpha-low beta or high beta range. Therefore, the pause-burst pattern of activity inside the STN contributes greatly to PD motor signs, and this is in accordance with the in vitro research performed by Beurrier et al. (1999) on rat brain sections (18). They found that about half of STN neurons may switch from tonic to burst firing mode when receiving inhibitory afferent projections, hyperpolarizing the cellular membrane. Moreover, activation of metabotropic glutamate receptors on the STN neurons may again favor switching to bursting mode.

Based on our findings, we may assume that synchronization between single units occurs more easily for bursting neurons, although this process affects all neurons in the STN motor area. Another possibility is that excessive synchronization affects predominantly pause-burst single units that have switched to this mode of action following the imbalance in the STN afferent projections under PD (Beurrier et al., 1999)(18). If this holds true, the prevalence of pause-burst neurons may be seen as an independent pathophysiological marker of PD, while excessive rhythmic synchronization of these units in the alpha-beta frequency range may represent the consequent stage of disease progression. Further research is necessary to examine this possibility.

## Data Availability

All data produced in the present study are available upon reasonable request to the authors

## Author contributions

Elena Belova: data collection, data pre-processing and statistical analysis, manuscript preparation and editing;

Veronika Filyushkina: data collection and data pre-processing;

Indiko Dzhalagoniia: data analysis;

Anna Gamaleya: neurological assessment of PD patients before the surgery;

Alexey Tomskiy: running DBS neurosurgeries;

W.-Julian Neumann: data interpretation, manuscript editing

Alexey Sedov: data collection, data pre-processing, manuscript editing, study management.

